# Taranto’s long shadow? Cancer mortality shows alarming peaks for specific types in the most polluted city of Italy but also in surrounding towns

**DOI:** 10.1101/2020.12.18.20248464

**Authors:** R. Roberto Cazzolla Gatti, Alena Velichevskaya

## Abstract

A national-scale study in Italy showed an incidence of cancer higher in the territories indicated as highly polluted compared to the regional average. One of them, the city of Taranto in Apulia (Italy), which is considered one of the most polluted cities in Europe, has numerous industrial activities that impact population health. We studied the epidemiological effects of a high level of pollution produced by the industrial area of Taranto in increasing the mortality rate for some specific cancer types in the city and towns of the two provinces located downwind. We analysed 10-year mortality rates for 14 major types of tumours reported among the residents of Taranto, of 6 surrounding towns, randomly placed within an imaginary cone in the main wind direction from the vertex of the industrial zone of Taranto. Our results confirm our hypothesis that the mortality rate for some specific types of cancer (namely, Hodgkin and non-Hodgkin lymphomas, leukaemia, liver and bladder tumours) are higher than the norm in the municipality of Taranto and we have evidence that other local causes may be implicated in the excess of mortality besides the potential dispersal of pollutants from the industrial area of Taranto. The proximity to the industrial area of Taranto cannot, therefore, explain alone the anomalies detected in some populations. It is likely that other site-specific sources of heavy pollution are playing a role in worsening the death toll of these towns and this must be taken into serious consideration by environmental policy-makers and local authorities.

## Introduction

Air pollution and, specifically, atmospheric particulate matter (i.e. also known as fine dust or PM) are catalogued among the confirmed carcinogens for humans [1]. Recent research and reviews of scientific studies on the subject determined that air pollution can cause lung cancer and increases the risk of bladder tumours [2-4]. One of the main sources of pollution is the industry, followed by urban traffic, agricultural emissions, heating, etc. [5]. A national-scale study in Italy showed an incidence of cancer (of various types) 9% higher in males and 7% higher in females compared to the regional average in the territories indicated as highly polluted [6].

In one of them, the city of Taranto in Apulia (Italy), there are numerous and diverse anthropogenic impacts connected to the presence of industrial activities. The city of Taranto has great importance for the Italian economy and is home to a large industrial, commercial and military port, the arsenal of the Italian Navy and an important industrial centre with steel plants (including Ilva, the largest steel centre of Europe), petrochemical (ENI refineries), cement (Cementir) and shipbuilding [7].

Multiple environmental monitoring studies and campaigns of measurement of industrial emissions carried out in the Taranto area highlighted a widespread environmental pollution situation and the significant contribution of the industrial centre, in particular the steel plant and the petrochemical complex, as a source of pollutants of sanitary interest [8].

The population of Taranto, together with that of the other main Italian cities included in the list of highly polluted areas, has been the subject to several epidemiological and health impact studies that have documented the role of air pollution on the increase of short and long term effects, such as mortality and morbidity for tumours [9].

The presence of critical issues regarding the pathologies associated with the pollutants emitted by the factories in the industrial area of Taranto, confirmed by the local health agency [10], induced many institutions and organization to claim the closure order of the factories in the territory. However, the industrial area has remained largely operative during the last decade. At the same time, the excess of tumour pathologies, with particular reference to lymphopoietic cancers, has kept a high death toll [11]. Another study to assess the exposure of Taranto citizens to dioxins and PCBs through the analysis of breast milk confirmed that, on average, the concentration of these substances in women residing in Taranto was 28% higher than those residing in the province and in line with what observed in other industrialized areas in Italy [12].

The effects of air pollution dispersal in increasing the cancer mortality rate of surrounding populations from the main industrial source (such as that of Taranto) have been, however, poorly explored [13] and mainly focused on “wind-days” [14]. Although it is well-known that air pollutants can disperse over long ranges, affecting also the locations that are far from the source and not only those in its extreme proximity, very few studies have investigated this issue and found contrasting results [see for instance 15-16].

Here we hypothesize that the high level of air pollution emitted by the industrial area of Taranto increases the mortality rate for some specific cancer types in the city but also impacts the towns of the two provinces located downwind. To test our hypothesis we analysed 5-year mortality rates for 14 major types of tumours reported among the residents of Taranto, of 6 surrounding towns - randomly placed within an imaginary cone in the main wind direction from the vertex of the industrial zone of Taranto -, and the two provinces (Bari and Taranto) to which these municipalities belong to.

## Methods

Considering that winter winds blowing in the Gulf of Taranto from N-NO disperse most of the atmospheric pollutants from the city of Taranto to the Ionic sea, to analyse the potential epidemiological effects of air pollution dispersion from a selected source (the industrial area of Taranto) to inhabited surrounding areas, in this study we considered the main direction and frequency of winds blowing towards mainland [17]. These are spring and summer SO breezes and SE winds in autumn (see the inset of wind directions in Fig. 1). On an annual basis, the direction N has the greatest persistence (75 hours with 16 knots of average speed), followed by S direction (69 hours with 13 knots of average speed) and E (66 hours with 9 knots of average speed). The average direction of winds interesting for our analysis was S-SE. As study sites we, therefore, randomly selected 6 municipalities, at different distances (0-60 km), within an imaginary cone of pollutants dispersion in direction S-SE from Taranto (as the vertex of the imaginary cone; Fig. 1). We also included the city of Taranto as the 7^th^ study site and the two provincial areas in the region that may potentially be most affected by the epidemiological consequences of air pollution from the industrial area of Taranto: the Metropolitan Areas of Bari and Taranto.

**Figure 1.**
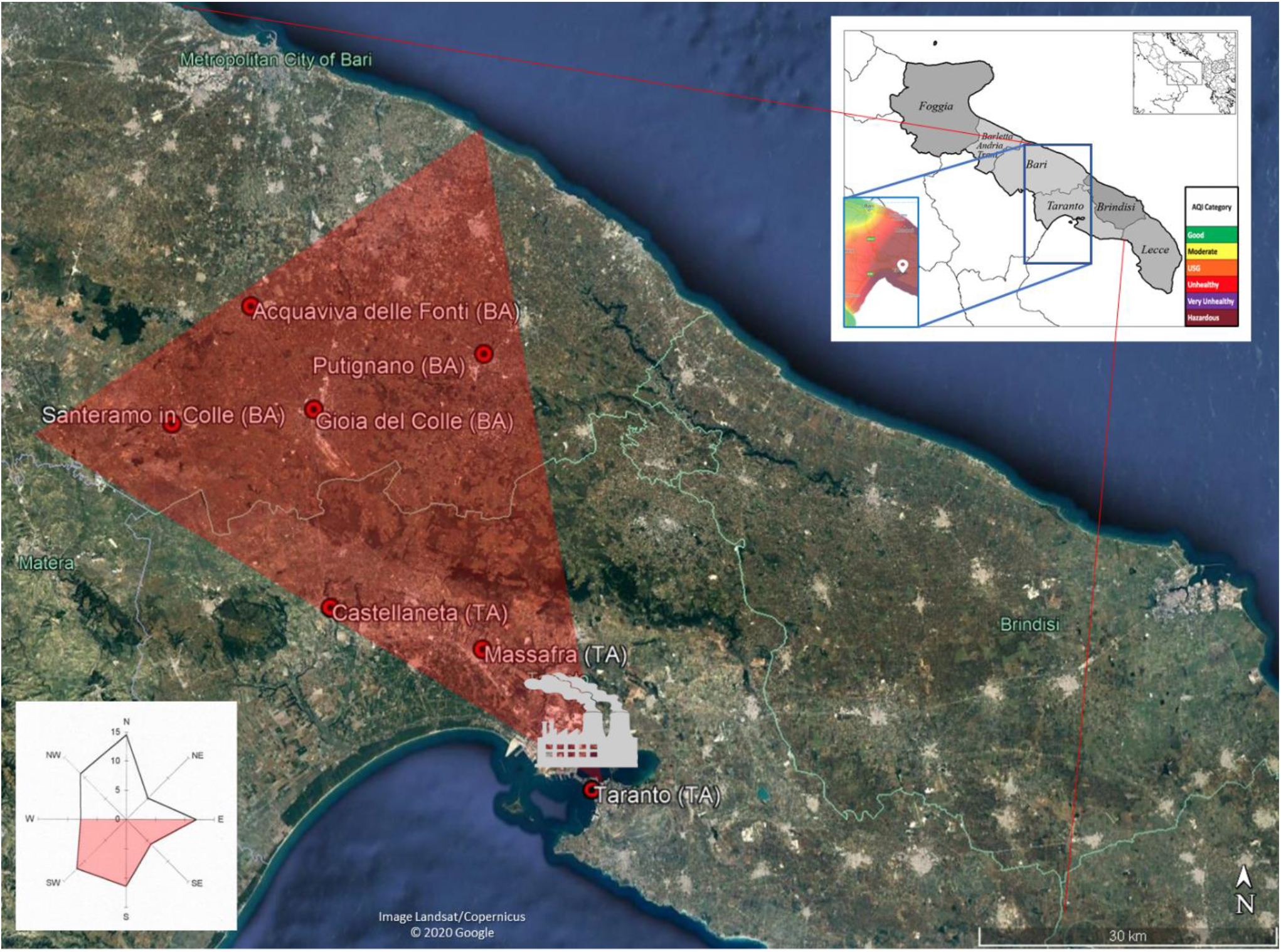
The location of the 7 study sites in an imaginary cone of the wind directions towards the mainland. The lower-left inset shows the frequency of the wind direction and, highlighted in red, are those directed towards inhabited areas. The red imaginary cone on the map with the industrial area of Taranto as the dispersion vertex of pollutants was placed to randomly select 6 municipalities, besides Taranto, in S-SE wind direction. The Provinces of Bari and Taranto, the two potentially most affected provinces in the region, are shown in the upper-right inset with a representative heat map that shows the Air Quality Index (AQI; green: low pollution, brown: high pollution) in Taranto and surrounding areas in an average day

Epidemiological data provided by the Italian National Institute of Statistics (ISTAT; [18]) of the resident population from 2012 to 2016 of the most recent five-year period (available at the time of data collection) in Italy, for the provinces of Bari and Taranto and the territory of the 7 municipalities above mentioned were collected for this study. Data on mortality causes divided by age groups (0; 1-4; 5-9; 10-14; …; 95+) were collected at national level (Italy). Mortality referred to all form of cancer were extracted from the datasets. For the additional analysis of the time series of blood cancers (lymphomas and leukaemia), annual epidemiological data from 2007 to 2016, at the provincial and municipal level and death per age groups (0; 1-4; 5-9; 10-14;…; 95+) at the national level, were collected.

The analysis was carried out using the statistical software R [19] on datasets in csv format.

To carry out a statistical comparison between the different resident populations, neutralizing the effects deriving from their different age structures, the results were further standardized through the use of the indirect method [20]. The indirect method, defined as the Standardized Mortality Rate (SMR), is based on the ratio between the deaths observed in a territory and those expected in the same. The expected deaths were calculated by applying the corresponding specific mortality ratios of the population assumed as standard (the national one in this analysis) to the average annual population by age classes of each territorial unit. The SMR, therefore, expresses the relationship between the deaths observed in a specific territory and the expected deaths if in the same territory there was the annual mortality, specific for age groups, of the population used as standard. The confidence intervals (C.I.) of the standardized mortality ratios were calculated at 90% using the Poisson model, providing an upper and lower bound. The 90% C.I. lower bound was used as the reference to define the significance level of the excess of mortality compared to the national standard.

Regression analysis and the estimation of the Pearson’s correlation coefficient (r) was conducted for the 10-year time series of lymphomas and leukaemia SMR for the municipalities of Taranto (TA), Massafra (TA) and Gioia del Colle (BA) – those study areas that showed a constant over the standard or increasing mortality rate in 2012-2016 compared to 2007-2011 - and the Provinces of Bari and Taranto for comparison.

## Results

### Standardized mortality rate for all tumour types

From the analysis of the standardized mortality rate (SMR) for all tumours (i.e. from all the causes of cancer death recorded throughout the national territory), between 2012 and 2016, mortality in the municipality of Taranto resulted above the national level only for benign cancers and those of uncertain behaviour whereas the 90% C.I. lower bound of malignant and all tumours were close to the Italian standard value (Fig. 2).

**Figure 2.**
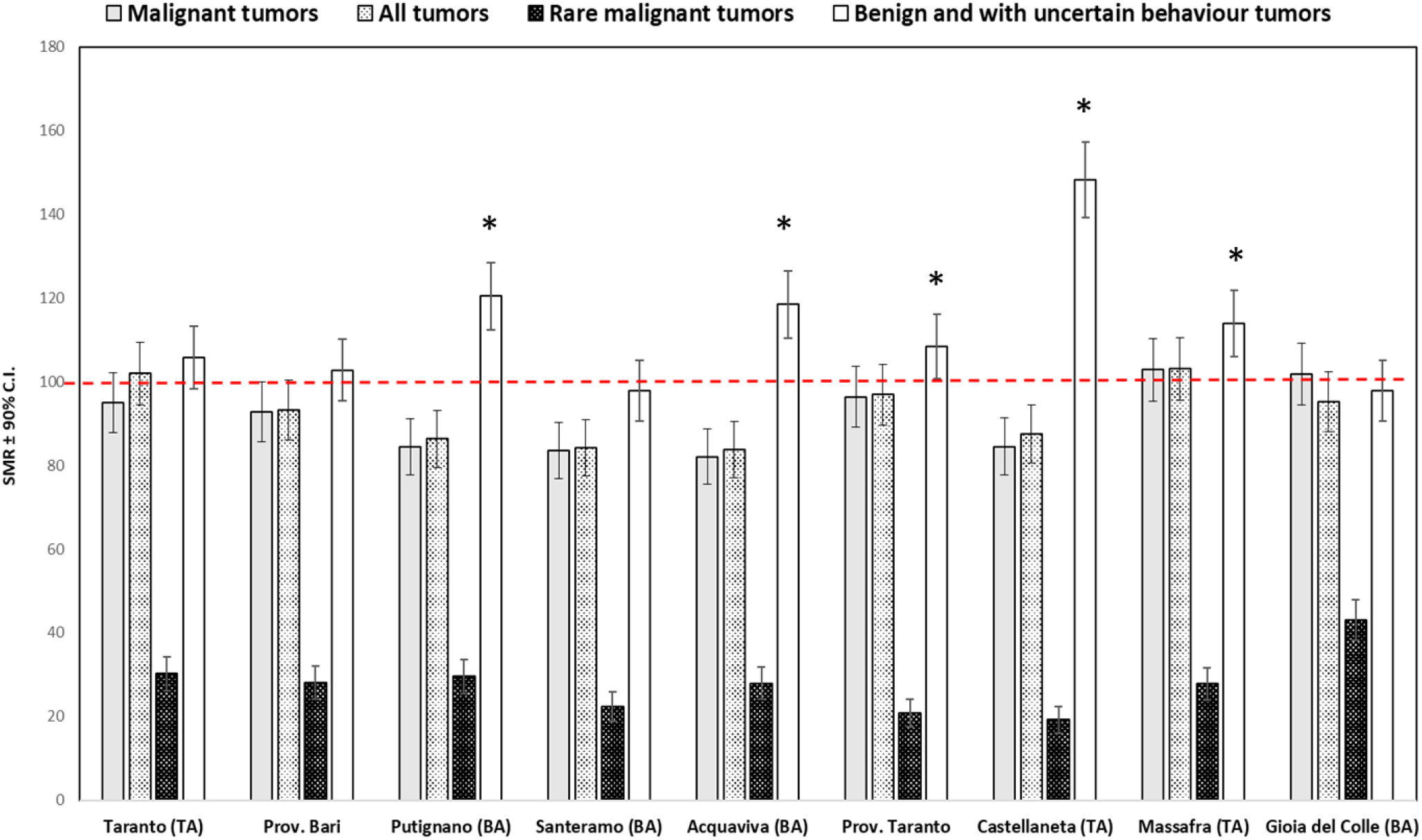
The 2012-2016 standardized mortality rate (SMR) in the population of the study sites for all, malignant only, benign and of uncertain causes, and rare tumours. Buffers indicate 90% C.I. The dashed horizontal line at 100 shows the national standard rate. Asterix indicates the statistical significance (above the 90% C.I. lower bound)

The municipalities of Putignano (BA), Acquaviva delle Fonti (BA), Castellaneta (TA), Massafra (TA) and the whole Province of Taranto show mortality for benign cancers and those of uncertain behaviour that fairly exceeds the national level. None of the study sites shows mortality for all and malignant tumours higher than the national level, although the municipalities of Massafra (TA) and Gioia del Colle (BA) show higher mortality for malignant tumours than surrounding areas, slightly above the national standard on average but within the 90% C.I. Moreover, the municipality of Massafra (TA) also shows mortality for all tumours slightly above the national standard on average but within the 90% C.I. (Fig. 2)

Because in the case of rare tumours, the types of all cancers that occur at the local/municipal level is unlikely to include the entire vast set of those that occur throughout the reference country, it seems - therefore - significant to notice that the municipality of Gioia del Colle (BA), although fairly below the national level, shows a mortality rate for rare tumours significantly above all the other municipalities (including Taranto) and both the Provinces of Bari and Taranto (Fig. 2).

From the analysis of the standardized mortality rate (SMR) of all the specific types of malignant tumours (i.e. all the causes of cancer reported at the national level) in the five years 2012-2016, more details emerge.

Mortality for liver and intrahepatic bile duct tumours among the residents of the municipalities of Taranto (TA), Santeramo (BA), Massafra (TA) and Gioia del Colle (BA) and the Provinces of Bari and Taranto are significantly higher than the national average (Fig. 3a). The mortality rate of liver and intrahepatic bile duct tumours of the municipality of Gioia del Colle (BA) highly exceeds the national level, that of all the other municipalities and the two provinces analysed. Only the municipalities of Taranto (TA) and Massafra (TA) shows mortality for all stomach tumours slightly above the national standard on average but within the 90% C.I. (Figure 3a). The municipalities of Castellaneta (TA) and Massafra (TA) show mortality for colon, rectus and anus tumours significantly higher than the national level, while the municipality of Gioia del Colle (BA) shows a rate slightly above the national standard on average but within the 90% C.I. (Figure 3a). None of the study areas exceeds significantly the national mortality rate for pancreatic cancer, although the municipality of Massafra (TA) shows a rate slightly above the national standard on average but within the 90% C.I. (Figure 3a).

**Figure 3.**
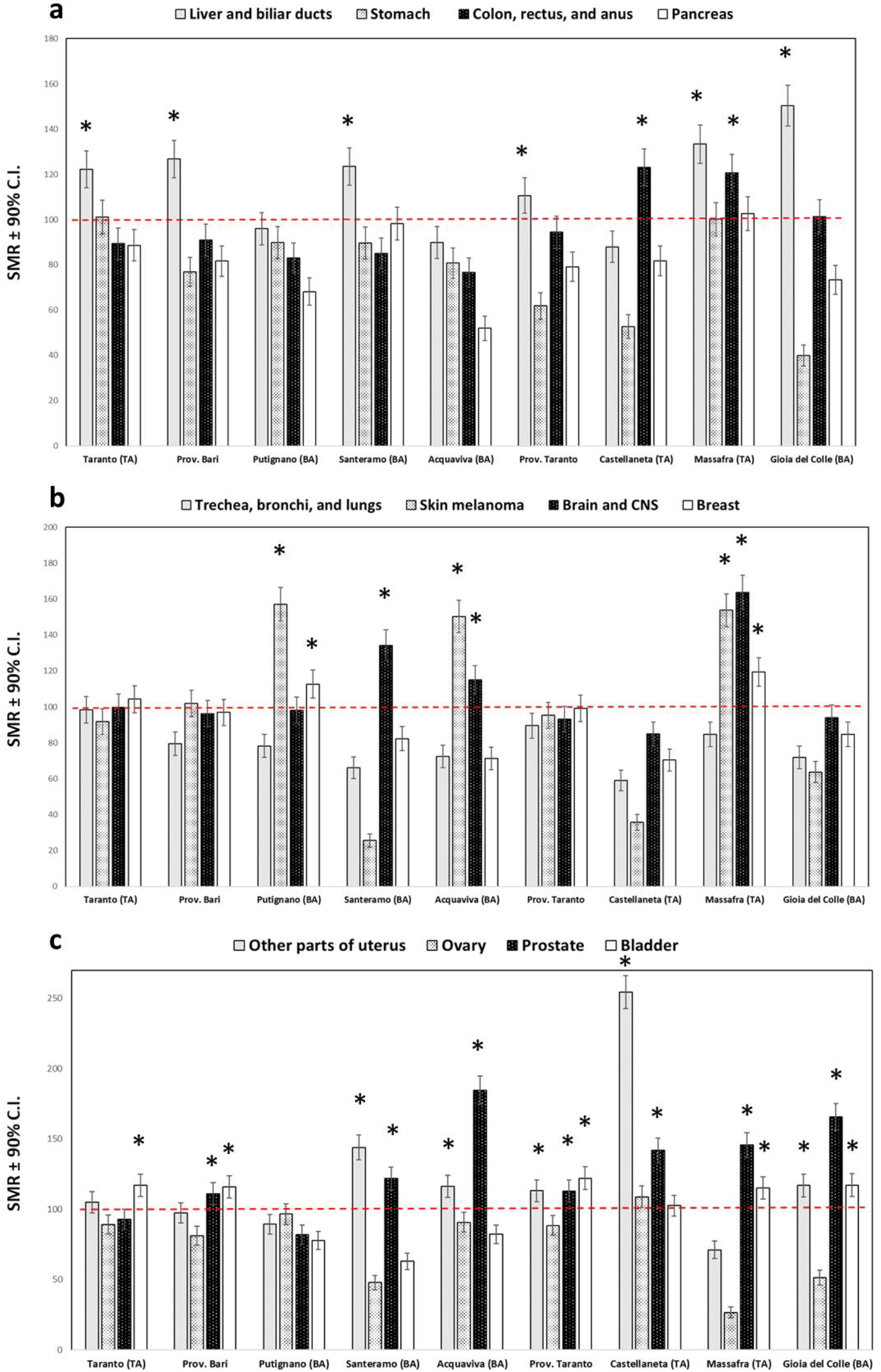
The 2012-2016 standardized mortality rate (SMR) in the population of the study sites for recorded types of tumours. Buffers indicate 90% C.I. The dashed horizontal line at 100 shows the national standard rate. Asterix indicates the statistical significance (above the 90% C.I. lower bound)

Mortality for trachea, bronchi, and lung tumours is not significantly higher than the standard in none of the study sites (Fig. 3b). Skin melanoma has an SMR exceed the standard in the municipalities of Putignano (BA), Acquaviva (BA), and Massafra (TA) (Fig. 3b). Mortality for brain and Central Nervous System (CNS) tumours is significantly higher in the municipality of Santeramo (BA), Acquaviva (BA) and Massafra (TA) (Fig. 3b). Only Putignano (BA) and Massafra (TA) show an excess of mortality for breast tumours, although the municipality of Taranto (TA) shows a rate slightly above the national standard on average but within the 90% C.I. (Figure 3b).

Ovary cancer mortality is above the standard only in the municipality of Castellaneta (TA), which shows even higher mortality for tumours in the other parts of the uterus (Fig. 3c). The municipalities of Santeramo (BA), Acquaviva (BA) and Gioia del Colle (BA) and the Province of Taranto also show an excess of mortality for this latter type of cancer (Fig. 3c). All municipalities, except for those of Taranto (TA) and Putignano (BA) show an excess of mortality for prostatic cancer, with the rate of Acquaviva (BA) and Gioia del Colle (BA) above all the others (Fig. 3c). Mortality for bladder cancer is higher than the standard among the residents of Taranto (TA), Massafra (TA), and Gioia del Colle (TA) and the population of the Province of Bari and Taranto (Fig. 3c). In the study period 2012-2016, there have been no recorded cases of cancer mortality (with n°≥5) for the lips, oral cavity, and pharynx, oesophagus, larynx, cervix, kidney and thyroid gland, in all municipalities.

### Standardized mortality rate for lymphomas and leukaemia

We then focused on the calculation of the standardized mortality rate (SMR) for blood cancers: lymphomas and leukaemia. We found that the municipality of Taranto (TA), Massafra (TA), Acquaviva delle Fonti (BA) and Gioia del Colle (BA) and the Province of Bari and Taranto recorded a mortality rate for lymphomas and leukaemia higher than the national reference level in the five years 2012-2016 (Figure 4). This was also significantly above all the other study sites in the residents of Massafra (TA) and Gioia del Colle (BA), in particular.

**Figure 4.**
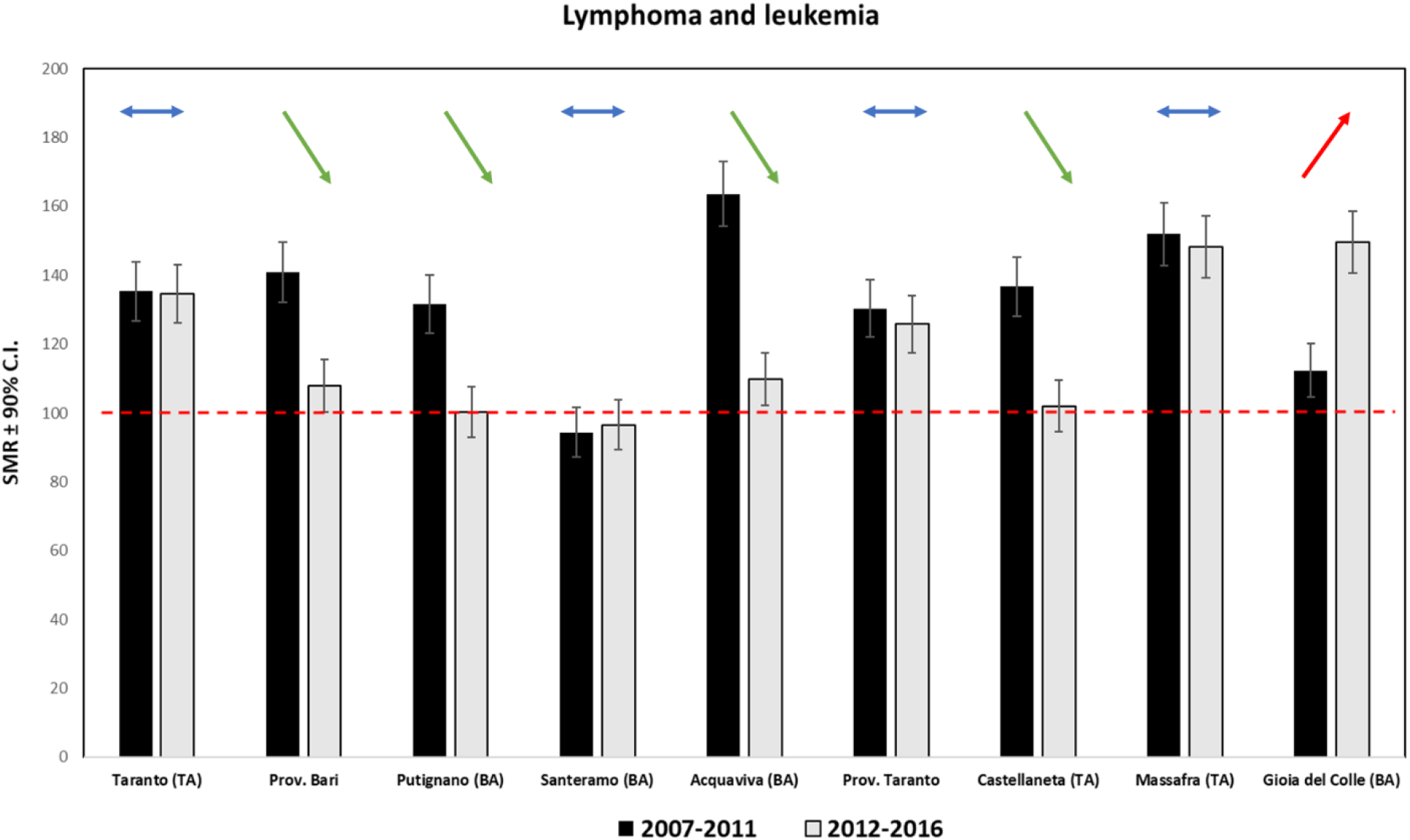
The comparison between the 2007-2011 and 2012-2016 standardized mortality rate (SMR) in the population of the study sites for blood cancers (lymphomas and leukaemia). Buffers indicate 90% C.I. The dashed horizontal line at 100 shows the national standard rate. Arrows show the significance at the 90% C.I. lower bound of the 5-to-5 year trend (red: positive, green: negative, and blue: stable)

To better understand the mortality rate dynamics in the study site for these two important types of tumours, which are commonly related to pollution effects on the human body [21, 22], we analysed the SMR for the previous 5-years period (2007-2011) for comparison. It emerged that, except for Santeramo in Colle (BA), all the municipalities and both provinces had an exceeding rate of mortality for lymphoma and leukaemia from 2007 to 2011 compared to the national level. Then, in the next 5 years (2012-2016), it strongly decreased in almost all study sites but kept quite constant in Taranto (TA), Massafra (TA), and the Province of Taranto (TA) and increased only in the municipality of Gioia del Colle (BA) (Fig. 4).

From a more detailed evaluation of the number of deaths observed in the only study site with an increasing trend, we found that Gioia del Colle (BA) had a mortality rate for lymphomas and leukaemia higher than the expected values every year of the period 2012-2016, with particularly high rates for lymphomas in 2016 (181% more mortality than expected) and for leukaemia in 2015 (111% more mortality than expected). From the comparative analysis with the decade 2007-2016, it emerged that the total mortality rate for leukaemia and lymphomas in the five years 2012-2016 increased of about 40% among the residents of Gioia del Colle (BA) (Fig. 4).

In an attempt to clarify these results, we also analysed the trend of the standardized mortality rate for lymphomas and leukaemia in the decade 2007-2016 for those municipalities that did not show a significant decrease below the standard in the two of 5-year periods of comparison (Fig. 4), namely Taranto (TA) and Massafra (TA), and of the only municipality which showed a significative increase, namely Gioia del Colle (BA). The 10-year time series for Hodgkin and non-Hodgkin lymphomas showed a slight increase for Taranto (TA) (slope=2.23, r=0.24), a steeper trend for Massafra (TA) (slope=11.51, r=0.6) and a highly steep trend for Gioia del Colle (BA) (slope=20.85, r=0.83). In Gioia del Colle (BA) there was a constant average increase that, between 2010 and 2012, exceeded the national average level and approached a 300% excess of mortality (approximately 181.44% of cases observed compared to those expected) around 2016 (Fig. 5a). This figure appears diametrically opposite to the trends of mortality from lymphomas and Hodgkin’s disease of both the provinces of Bari and Taranto in the decade 2007-2016, which showed a decreasing tendency over time that aligns with the national average value in the Province of Bari and is, on average, always below the standard in the Province of Taranto (Figure 5b). From 2007, the trend of leukaemia mortality decreased for the municipality of Massafra (TA) and Gioia del Colle (BA), although remained slightly above (on average) the national level during the 10 years, but was constant and always higher than the national standard among the residents of the municipality of Taranto (TA) (Fig. 5c). Trends in two provinces of Bari and Taranto decreased between 2007-2016 but remained above the national rate (Fig. 5d).

**Figure 5.**
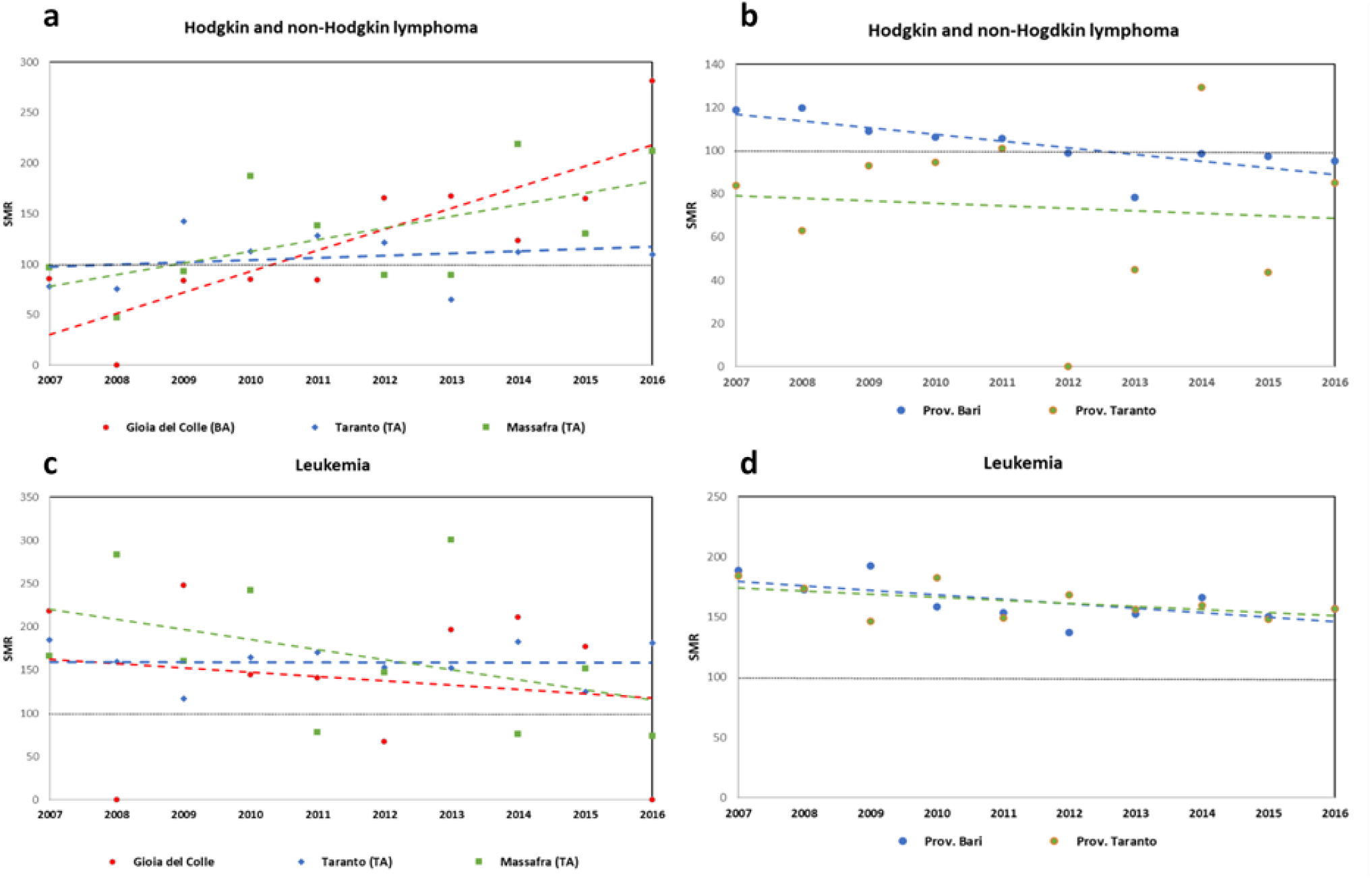
Mortality trends from 2007 to 2016 in the population of the municipalities of Taranto (TA), Massafra (TA), and Gioia del Colle (BA) for a) lymphomas and c) leukaemia. The same trends are shown for the Provinces of Bari and Taranto for b) lymphomas and d) leukaemia. Dashed horizontal line at 100 show the national standard rate.

## Discussion

According to the World Health Organization (WHO; [23]) and the main cancer research institutes [1], in addition to a genetic component, often unknown, there are well-known environmental factors that increase the risk of developing certain types of tumours and, in particular, lymphomas and leukaemia, such as ionizing radiation and exposure to chemical-industrial substances, agricultural pesticides, and products derived from benzene.

Although the mortality for the overall malignant and benign cancer types in the city of Taranto is close to the national rate, a more detailed analysis revealed interesting insights. The results of this epidemiological study, in fact, dramatically confirm our hypothesis that the mortality rate for some specific types of cancer (namely, Hodgkin and non-Hodgkin lymphomas, leukaemia, liver and bladder tumours) are higher than the norm in the municipality of Taranto, which is directly exposed to the pollution produced in the industrial area of the city.

Our overall result is that residents of the Provinces of Bari and Taranto are, in general, more affected by mortality due to leukaemia, prostate, liver, and bladder tumours than the average Italian citizens. Residents of Taranto show excessive mortality for 4 out of 14 cancer types (including high rates for lymphomas and leukaemia). However, we have evidence that residents of our study sites who live closer to Taranto (i.e. Castellaneta and Massafra) have differential rates for each specific cancer type. This suggests that other local causes may be implicated in the excess of mortality, besides the potential dispersal of pollutants from the industrial area of Taranto at a downwind distance. In fact, Massafra – which is only ∼13 km linearly far from the industrial area of Taranto - shows an excess of mortality for most of the reported causes of tumours (9 out of 14, including all the 4 types detected in Taranto, plus additional 5), much higher than Taranto. Among all municipal and provincial rates analysed, the epidemiological situation of Massafra (TA) for most cancer types appears dramatic. Castellaneta - which is just ∼14 km farther from Taranto on a linear distance – shows lower excess (4 out of 14 cancer types) than Massafra, similar in number to that of Taranto (although different in types: colon, uterus, prostate, and ovary).

Moving along linear distance in the dispersion cone we drafted on the map for this research (Fig. 1), we find the municipalities of Santeramo in Colle, Gioia del Colle and Putignano placed at about 40-50 km in the Province of Bari, almost equidistant from the industrial area of Taranto on an east-west fan-shaped location.

Putignano, with excesses only in skin melanoma and breast tumours (2 out of 14 cancer types) resulted in the municipality with relatively least concern among all. Equidistantly, residents of Santeramo showed higher mortality than national rates only for liver, brain, uterus, and prostate (4 out of 14). Gioia del Colle, after Massafra, represents – instead - the second-highest concern municipality with 6 out of 14 cancers exceeding the national standard mortality (liver, uterus, prostate, and bladder plus an enhanced rate of mortality for leukaemia and a constantly growing rate for Hodgkin and non-Hodgkin lymphomas).

Acquaviva delle Fonti, which is the farthest town from the industrial area of Taranto among our study sites (5 km northern than Gioia del Colle in a linear direction), shows exceeding rates for 5 out of 14 tumour types: melanoma, brain, uterus, prostate, and leukaemia.

From our results, we can, therefore, identify some common patterns. Those cancers rates that are in excess in the city of Taranto are also higher in the municipalities of Gioia del Colle and Massafra. Only Putignano, among all study sites, shows a low rate of cancer mortality for most of the specific tumour types. Besides the confirmation that some types of tumours are higher than the norm in the municipality of Taranto, it is noticeable that residents of Massafra (TA) and Gioia del Colle (BA) show higher tumour mortality rates that appear to be site-specific.

Besides the downwind position from Taranto’s industrial area, apparently, there might be other sources of pollution which could explain such high mortality rates in these two towns. In fact, many citizens of Massafra and Gioia del Colle, in recent years, manifested apprehension regarding the possible dispersion of pollutants from the waste incinerator of Massafra [24] and the experimentation plants on flameless and waste combustion (DISMO and CCA managed by ITEA and Ansaldo; [26]) located a few hundred meters from the houses in the two municipalities.

The town of Massafra (TA), because of its proximity to Taranto, in the 90s was included in the “area at high risk of environmental crisis”, as established by a decree of the Council of Italian Ministers [27]. Despite this, in 2003 the first plant for the production of electricity from fuel derived from waste incineration (CDR, a plant managed by Appia Energia Srl) came into operation. On December 16, 2013, the Regional Environmental Protection Agency (ARPA Puglia), in an information note sent to all the local administrative authorities, including the mayor of Massafra, announced that the value of the maximum concentration of dioxins and PCBs exceeded in a sample of bovine milk collected from a farm located in the countryside of the town. Subsequent confirmatory analyses of additional milk samples showed an even more unfavourable outcome about PCDD/F and PCB contamination. The same regional agency (ARPA Puglia) in 2017 verified that the incinerator manager did not carry out any monitoring campaign for PM10, PM2.5, metals, and organic pollutants. Yet, during the inspection of soil samples (on agricultural land) a concentration of beryllium and vanadium above the threshold for metals, which are carcinogenic for human beings was found [28]. Then, ARPA reported that even the company’s self-checks in 2014 and 2017 had shown an excess above the limits for beryllium and also for tin [28].

In Gioia del Colle (BA), experiments on waste treatment have been conducted for decades - during and before the period considered in this study - by ITEA and Ansaldo on a flameless plant called DISMO and on an incinerator plant called CCA, with the combustion of various types of waste, among which special and radioactive wastes [29].

In 2019 authorities acted a preventive seizure of the plant because of the “documented illegal disposal of dangerous waste containing carcinogenic and mutagenic of the maternal foetus chemical compounds, acid sludge, and residues of industrial chemical reactions” [30]. In addition, the extreme proximity to the inhabited centre of the city of Gioia del Colle of the “36th Storm” military airport, where radioactive weapons were stored [31], military radars are in operation, nuclear wastes deriving from the experiments carried out at the ITREC nuclear plant in Basilicata were delivered [32], and from which combat aircraft take off daily and can release huge quantities of exhaust gases that also contain carcinogens over the areas surrounding the town [33, 34], has always caused great concern in the local population.

From recent analyses carried out by ARPA Puglia [35, 36] on the air quality of the urban centre of Gioia del Colle, in fact, an indicative data concerning the toluene/benzene (T/B) ratio emerged. This ratio allows distinguishing the sources of chemical-industrial contamination from those of vehicular-road air pollution [37, 38]. Values of this ratio above 4.5 and below 1.5 were detected in over 70% of the monitoring days in the municipal area, with high fluctuation in the sampling period, between December 2018 and June 2019 (i.e. with fluctuations recorded mainly on weekdays and seldom on holidays). These values could be indicative of an intermittent source of chemical-industrial pollution, such as a factory that works on alternate days (an experiment on waste incinerator?) or an airport operating mainly on selected working days (a military airport?). Toluene, in fact, is a relevant pollutant of factory productions [39] and a component military jet fuels, as it can increase the octane number and, therefore, the power of the aircraft’s engine [40].

It is also interesting to notice that from the analysis of the standardized mortality rate (SMR), rare malignant tumours (i.e. the set of malignant tumours that do not fall into the categories of frequent ones at national scale) among the residents of Gioia del Colle are much higher compared to the rate of all the other municipalities and the two provinces considered in this study (Fig. 2).

Most of our study sites, including the low concern town of Putignano, showed exceeding mortality rates for common cancers, such as breast, skin, uterus, and prostate. Differently, mortality for less common tumours such as those of liver and bladder was particularly high among the residents of Taranto, Massafra, and, above all, Gioia del Colle. Among the well-known environmental causes of liver and intrahepatic bile ducts tumours, there is the exposure to fine particles PM2.5 [41], various industrial chemicals [3, 42], and ionizing radiation [3, 42]. Furthermore, the high mortality from bladder tumours can be caused by pollution made of aromatic amines, compounds of industrial origin (which are released by combustion or by the treatment, for example, of tires, paints, and PVC) and pesticides, recognized environmental risk factors both for bladder and liver tumours [43-45].

## Conclusion

The concomitant and excessive mortality among the residents of Taranto, Massafra and Gioia del Colle for blood tumours (leukaemia and, particularly, lymphomas and Hodgkin’s disease) and for liver and bladder cancers suggests the presence, on the local territory, of significant sources of chemical-industrial and/or radioactive pollution such as to greatly increase the rates concerning both the national level and that of many neighbouring municipalities and the two provinces surveyed. The positive trend of mortality for lymphomas and Hodgkin’s disease in the decade 2007-2016 among the residents of Massafra and, even more, among those of Gioia del Colle compared to the opposite decreasing trend in the two provinces of Bari and Taranto, confirms the presence of one or more, constant over time, sources of contamination located in the municipal areas.

Similarly, the far higher mortality rate for rare tumours than in all the other municipalities and provinces considered, suggests the presence of localized environmental causes that favour, among the residents of Gioia del Colle, types of tumours that are recorded in a much lesser extent among citizens of the surrounding areas.

To shed more light on the problems that threat the environment and health and on the possible relationship between pollution and the incidence of some serious proliferative diseases affecting the citizens of Taranto, Massafra and Gioia del Colle, we recommend detailed medical and air quality monitoring (with analysis of chemical-industrial atmospheric pollutants such as IPA, Dioxins, PCBs, PM2.5, etc. and electromagnetic pollution), water (with analysis of the presence of heavy metals and radioactivity in drinking water) and soil (with analysis of chemical contaminants, pesticides, PAHs, PCBs, dioxins, heavy metals, etc. and radioactivity in soil and local food products).

In conclusion, we found that dispersion of air pollutants from the industrial area of Taranto can contaminate the surrounding environment, extending over kilometres of distance, and damage the health of people living in relatively distant localities, increasing their risk of mortality for some specific tumours. However, we discovered that some common and rare types of cancer have an uneven distribution in the provinces of Taranto and Bari. The proximity to the industrial area of Taranto cannot, therefore, explain alone the anomalies detected in some populations (particularly those of Gioia del Colle and Massafra). It is likely that other site-specific sources of heavy pollution are playing a role in worsening the death toll of these towns and this must be taken into serious consideration by environmental policy-makers and local authorities.

## Data Availability

All data used for this study are publicly available from ISTAT

https://www.istat.it/it/archivio/4216

## Author contribution statement

R.C.G. conceived the idea, analysed the data and wrote the manuscript. A.V. collected the data and support the analysis of them. R.C.G. and A.V. prepared figures and graphical material.

